# A comparison of four self-controlled study designs in an analysis of COVID-19 vaccines and Myocarditis using Five European Databases

**DOI:** 10.1101/2023.11.10.23298290

**Authors:** Anna Schultze, Ivonne Martin, Davide Messina, Sophie Bots, Svetlana Belitser, Juan José Carreras-Martínez, Elisa Correcher-Martinez, Arantxa Urchueguía-Fornes, Mar Martín-Pérez, Patricia García-Poza, Felipe Villalobos, Meritxell Pallejà-Millán, Carlo Alberto Bissacco, Elena Segundo, Patrick Souverein, Fabio Riefolo, Carlos E. Durán, Rosa Gini, Miriam Sturkenboom, Olaf Klungel, Ian Douglas

**Author notes:** Disclaimer: The research leading to these results was conducted as part of the activities of the EU PE&PV (Pharmacoepidemiology and Pharmacovigilance) Research Network which is a public academic partnership coordinated by the Utrecht University, The Netherlands. Scientific work for this project was coordinated by the University Medical Center Utrecht in collaboration with the Vaccine Monitoring Collaboration for Europe network (VAC4EU). The project has received support from the European Medicines Agency under the Framework service contract nr EMA/2018/23/PE. The content of this paper expresses the opinion of the authors and may not be understood or quoted as being made on behalf of or reflecting the position of the European Medicines Agency or one of its committees or working parties.

## Abstract

**Introduction:** The aim of this study was to assess the possible extent of bias due to violation of a core assumption (event-dependent exposures) when using self-controlled designs to analyse the association between COVID-19 vaccines and myocarditis.

**Methods:** We used data from five European databases (Spain: BIFAP, FISABIO VID, and SIDIAP; Italy: ARS-Tuscany; England: CPRD Aurum) converted to the ConcePTION Common Data Model. Individuals who experienced both myocarditis and were vaccinated against COVID-19 between 1 September 2020 and the end of data availability in each country were included. We compared a self-controlled risk interval study (SCRI) using a pre-vaccination control window, an SCRI using a post-vaccination control window, a standard SCCS and an extension of the SCCS designed to handle violations of the assumption of event-dependent exposures.

**Results:** We included 1,757 cases of myocarditis. In unadjusted analyses, agreement between study designs varied by vaccine brand. There was good agreement between all designs for AstraZeneca and Pfizer, but for Moderna we found harmful incidence rate ratios (IRR) using the standard and extended SCCS (standard SCCS: IRR = 3.12, 95%CI = 1.53 – 6.40; extended SCCS: IRR = 2.43, 95%CI = 1.11 – 5.33) compared with no association with the SCRIs (SCRI-pre: IRR = 0.60, 95%CI = 0.27 – 1.33; SCRI-post: IRR = 0.86, 95%CI = 0.34 – 2.19), although confidence intervals were wide. There was very good agreement between all designs for the unadjusted second dose analyses, confirming the known harmful association between the second dose of Moderna and Pfizer vaccines and myocarditis.

**Conclusions:** In the context of the known association between COVID-19 vaccines and myocarditis, we have demonstrated that two forms of SCRI and two forms of SCCS led to largely comparable results, possibly because of limited violation of the assumption of event-dependent exposures.

## Introduction

Self-controlled study designs are a useful tool for evaluating vaccine safety signals, as they do not require the identification of an external control group and automatically control for all time-invariant confounding (1). This is an important benefit when studying vaccinations, as those who choose to get vaccinated are often very different in terms of their underlying health from those who are unvaccinated (2). Two commonly used self-controlled designs in vaccine safety are the self-controlled case series (SCCS) and the self-controlled risk interval (SCRI) designs. In these study designs only people with the outcome (cases) are included, and the risk of an event occurring during a pre-specified risk window after the exposure is compared to the unexposed control period. In a typical SCCS, all time except the risk window is considered as the control period, that is, observation does not stop at the occurrence of the outcome. The SCRI is a special case of the SCCS in which the control period is fixed relative to the vaccination date (3).

Arguably the most important assumption for SCCS is that the occurrence of the outcome does not impact the probability of subsequent exposure (4,5). In the vaccine setting, this may be violated if future doses are delayed following the event of interest. For example, the occurrence of myocarditis may delay the receipt of a COVID-19 vaccine, resulting in an upward bias in the association between COVID-19 vaccines and myocarditis if ignored in analyses (6). A pre-exposure window can be used to correct for delays but will not suffice if the event delays the exposure for indeterminate periods of time or contraindicates it (7).

Alternative solutions include only analysing pre-event exposures and starting the observation time at the exposure (7), or using extensions of the SCCS based on a pseudo-likelihood approach (5). Starting the observation time at the exposure only works for a single vaccine dose, and simulation studies have demonstrated that it may introduce bias in the multi-dose setting (8). A less commonly used option involves truncating the SCCS observation time to only include a pre-specified time period between doses, during which another vaccination would not be expected to occur (9). This method is similar in its conceptualisation to an SCRI using one or more post-vaccination control windows.

Another important source of potential differences between SCCS and SCRI designs relates to calendar time trends: as the SCCS typically includes a longer study period, it may be more sensitive to variations in the occurrence of the outcome over time. However, the SCRI is theoretically more vulnerable to rapid changes in outcome incidence, as all comparisons are mono-directional. For example, an SCRI using a control period prior to a vaccine being administered might overestimate the risk of a safety event if the incidence of the safety event increases over calendar time.

The choice of study design is of key importance for studies of vaccine safety, but comparisons of versions of the SCRI and SCCS in settings where the assumption of event-dependent exposures may be violated are limited. We, therefore, compared these study designs using a case study of multidose COVID-19 vaccination and myocarditis, in which we were concerned that violation of the event-dependent exposure assumption, and therefore, the specification of the control periods, may be an issue.

## Methods

### Description of the SCCS and SCRI designs

All study designs were self-controlled, that is, they included only cases. In all designs, the risk period after each dose started on day 1 and lasted until day 28. The focus was on the first and second doses, as COVID-19 booster doses had not been rolled out at the time of design of this study. Where relevant, doses were assigned a pre-exposure period of 30 days to account for potential short violations of the assumption of event-dependent exposures.

The day of vaccination was modelled as a separate risk window. In the case of overlaps, risk periods always took precedence over pre-exposure periods, and latter risk periods took precedence over earlier risk periods. All designs are described in more detail below and illustrated in Figure 1.

a. Pre-vaccination SCRI The pre-vaccination SCRI used a 60-day period before the 1^st^ vaccine dose lasting from days [-89, -30] as control time. A 30-day period before each vaccine dose [-29, 0] was considered a pre-exposure period (Figure 1a). In practice, both pre-exposure periods and time in between doses, where this occurred, were treated as separate levels of the exposure variable. This means that individuals who experienced events in these time periods were included in the analysis, and will have contributed to the calendar time adjustment (although they did not directly contribute to the contrast of interest of risk versus control period). The estimates for those exposure levels are hard to interpret as the time contributed varied by individual, and these are therefore not reported.
b. Post-vaccination SCRI The post-vaccination SCRI used time after the second vaccine dose as control time, or time after the first vaccine dose for those without a second vaccine dose. A control period of 60 days was used, lasting from days [29, 88] after the relevant vaccine dose (the first 28 days constituted the risk period). The design is illustrated in Figure 1b below.
c. Standard SCCS The standard SCCS used all calendar time, from a fixed start date in calendar time (1 September 2020 until the last available follow-up (Figure 1c). The start of follow-up was set to provide enough time to implement a pre-vaccination SCRI before the receipt of the first vaccine in the contributing databases (8 December 2020. All time not designated as a pre-exposure period, a risk period or a vaccination day was used as the control period.
d. Extended SCCS This used the same design as the standard SCCS (Figure 1c) but with an analysis method based on a pseudo-likelihood approach. This method was developed specifically to account for bias that might be introduced due to event-dependent exposures (Farrington, Whitaker, and Hocine 2009).

**Figure 1.**
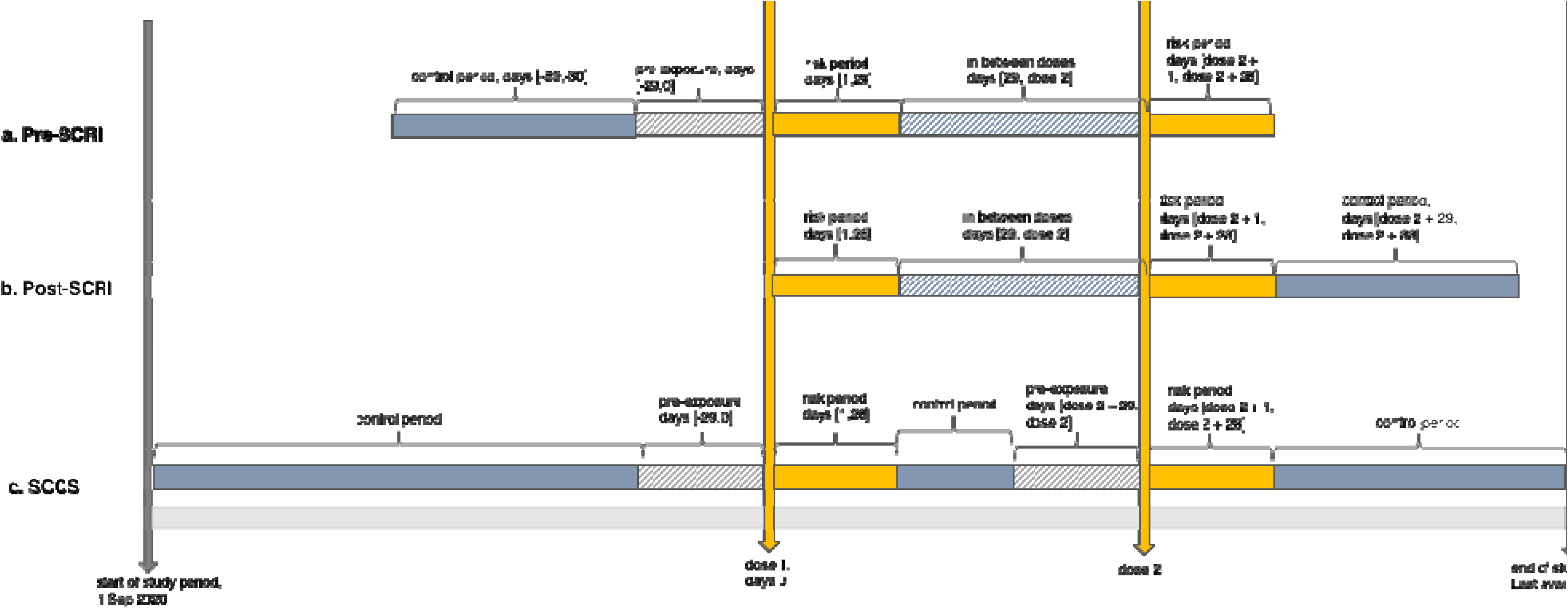
Illustration of the study designs

Our hypotheses regarding the extent of bias can be seen in Table 1 when the assumption of event-dependent exposures is violated.

**Table 1.**
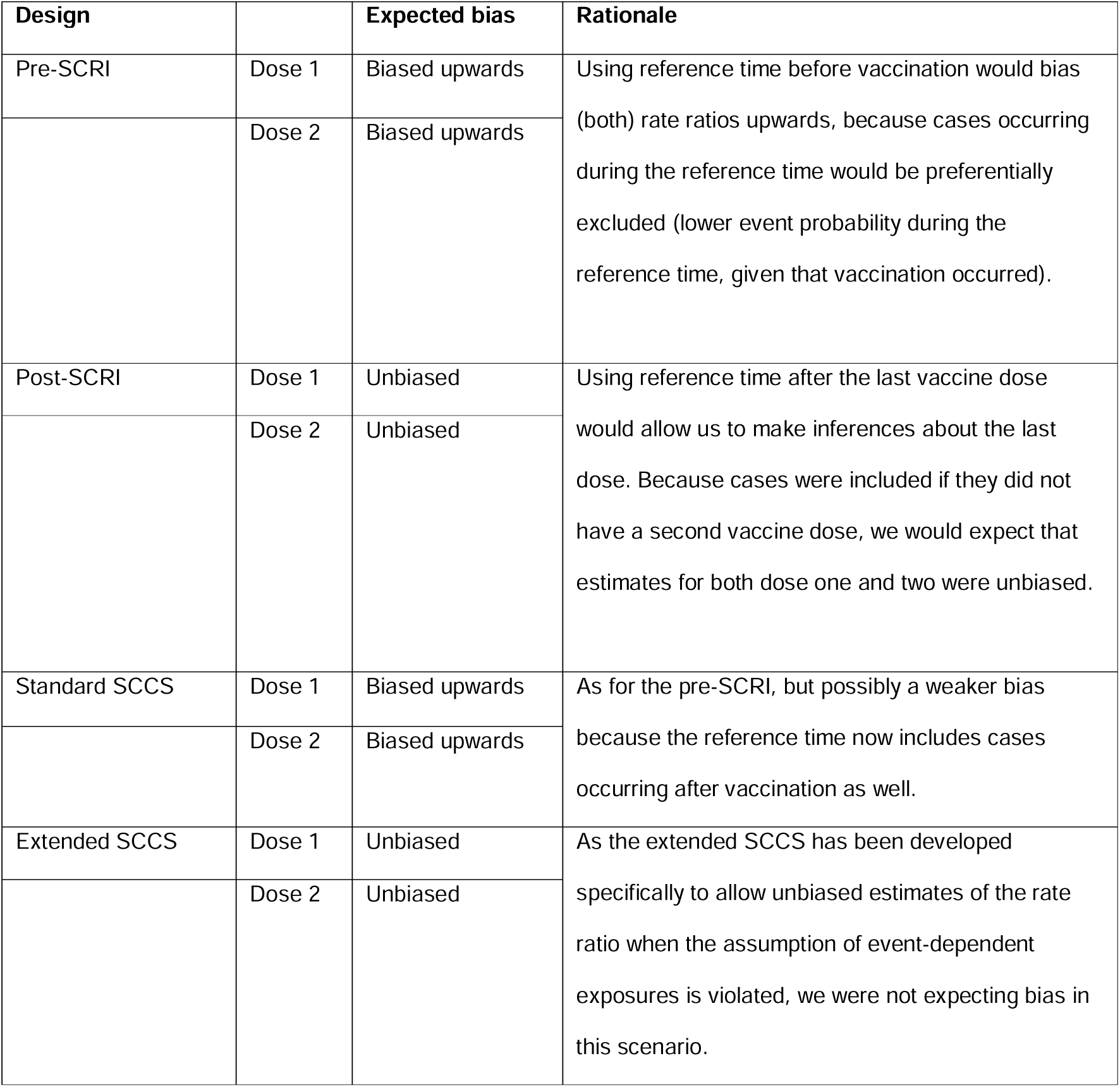
Expected bias under each design if the assumption of event-dependent exposures is violated.

| Design |  | Expected bias | Rationale |
| --- | --- | --- | --- |
| Pre-SCRI | Dose 1 | Biased upwards | Using reference time before vaccination would bias (both) rate ratios upwards, because cases occurring during the reference time would be preferentially excluded (lower event probability during the reference time, given that vaccination occurred). |
|  | Dose 2 | Biased upwards |  |
| Post-SCRI | Dose 1 | Unbiased | Using reference time after the last vaccine dose would allow us to make inferences about the last dose. Because cases were included if they did not have a second vaccine dose, we would expect that estimates for both dose one and two were unbiased. |
|  | Dose 2 | Unbiased |  |
| Standard SCCS | Dose 1 | Biased upwards | As for the pre-SCRI, but possibly a weaker bias because the reference time now includes cases occurring after vaccination as well. |
|  | Dose 2 | Biased upwards |  |
| Extended SCCS | Dose 1 | Unbiased | As the extended SCCS has been developed specifically to allow unbiased estimates of the rate ratio when the assumption of event-dependent exposures is violated, we were not expecting bias in this scenario. |
|  | Dose 2 | Unbiased |  |

### Case Study

We compared these designs as part of an evaluation of COVID-19 vaccination and myocarditis completed as part of the Covid Vaccine Monitoring (CVM) project, a collaboration between the EU PE&PV (Pharmacoepidemiology and Pharmacovigilance) Research Network (led by Utrecht University) and the Vaccine Monitoring Collaboration for Europe network (VAC4EU). The clinical findings using a pre-vaccination SCRI have been published (11).

#### Data

Data were included from the Spanish Base de Datos para la Investigación Farmacoepidemiológica en Atención Primaria (BIFAP) database (end of data; April 2022), the Italian ARS-Tuscany database (end of data; December 2021), the Spanish FISABIO VID database (end of data; December 2021), the Spanish SIDIAP database (end of data; June 2022) and the British Clinical Practice Research Datalink CPRD Aurum (end of data; March 2022). All five data providers converted their data to the ConcePTION Common Data Model which resulted in structurally harmonised local datasets (10), and have been described in greater detail in previous publications (11).

#### Study Design and Analysis

The core methods have been previously described (11). Briefly, individuals who experienced both the outcome of interest and were vaccinated between 1 September 2020 and the end of data availability (which varied in the different databases) were eligible for inclusion. We further required individuals to have at least one year of baseline time prior to the start of the study period, have non-missing age and sex, be aged 18 years or older at the start of the study period, have a known vaccine brand for their first dose, and no history of myocarditis in the 365 days leading up to the start of the study period. The outcome of interest was the first code of myocarditis during the study period (12). The vaccines of interest were Comirnaty (Pfizer/BioNtech), Spikevax (Moderna), Vaxzevria (AstraZeneca) and the Jcovden COVID-19 vaccine (Janssen). Due to small numbers and because our interest was primarily in the comparison of the different designs, we excluded individuals who received different brands of vaccine for their first and second vaccine doses.

Third and fourth doses were not considered as exposures of interest in this analysis, as the number of booster vaccinations in the data was very low at the time of the design. Nevertheless, the presence of further doses needed to be accounted for in the empirical analysis. How this was handled varied depending on the design. In the post-vaccination SCRI, we curtailed the control period at the start of the third dose, if this occurred. We expected such instances to be relatively rare given the minimum time required between second and booster doses but nevertheless quantified the occurrence of such curtailment in each database. For the standard and extended SCCS, we considered third and fourth doses, and their pre-exposure periods, as separate levels of the exposure variable to remove periods of potential increased risk from the reference time.

For each study design, we generated basic summary data for the people included in each analysis, plotted histograms showing the time between event and each vaccination (exposure-centred interval plots1), and fit conditional Poisson regression models to calculate incidence rate ratios for the association between the first and second vaccine dose and myocarditis, stratified by DAP and vaccine brand using the *{SCCS}* package in R. Finally, we added a random-effect meta-analysis across the data sources using the *{meta}* package in R.

Agreement between designs was assessed informally, by considering what the interpretation of findings would be from a public health perspective. All code for preparing the data and running these models is available on Github (note: link to be made public on paper acceptance; peer-reviewers can be given access on request).

## Results

### Description of Study Population

In total, we included 1,757 cases of myocarditis: 191 cases from ARS, 642 from CPRD, 240 from FISABIO, 404 from SIDIAP and 280 from BIFAP-PC. A flowchart showing the selection of individuals from each country is provided in Table 2.

**Table 2.** Selection into the case series, by DAP.

|  | ARS | CPRD | FISABIO | SIDIAP | BIFAP-PC |
| --- | --- | --- | --- | --- | --- |
|  | N (%) | N (%) | N (%) | N(%) | N(%) |
| Total individuals in dataset | 3704289 | 15214165 | 5607181 | 6220172 | 12912064 |
| non-missing gender | 3704289 (100) | 15214165 (100) | 5607181 (100) | 6220172 (100) | 12912064 (100) |
| non-missing age | 3704289 (100) | 15214165 (100) | 5607181 (100) | 6220172 (100) | 12912064 (100) |
| age over 18 | 3083301 (83.24) | 11682705 (76.79) | 4522073 (80.65) | 4995184 (80.31) | 10462443 (80.31) |
| non-missing first dose | 2527061 (68.22) | 7652699 (50.3) | 3715022 (66.25) | 4066912 (65.38) | 8402992 (65.38) |
| non-missing type of first dose | 2527061 (68.22) | 7652699 (50.3) | 3715022 (66.25) | 4066912 (65.38) | 8402992 (65.38) |
| dose 1 and 2 the same brand | 2436048 (65.76) | 7459138 (49.03) | 3527140 (62.9) | 3776589 (60.72) | 7946432 (60.72) |
| non-missing outcome | 511 (0.01) | 1295 (0.01) | 763 (0.01) | 738 (0.01) | 603 (0.01) |
| outcome occurs after start | 233 (0.01) | [redacted] | 281 (0.01) | 404 (0.01) | 280 (0.01) |
| outcome occurs before censor | 191 (0.01) | 642 (0) | 240 (0) | 404 (0.01) | 280 (0.01) |
| **Cases included** | **191 (0.01)** | **642 (0)** | **240 (0)** | **404 (0.01)** | **280 (0.01)** |

Most cases received the Pfizer/BioNtech vaccine (Table 3). The second dose coverage was around 80% or higher for all vaccine brands in all countries, although the number of third doses was relatively low for all brands. The post-vaccine SCRI, as planned, required a minimum of 89 days between the second and third vaccine dose (28 day risk period, plus a 60 day control period), and it was reassuring that the minimum time between the second and third dose was longer than 90 days for all brands in all countries, apart from for Moderna in the FISABIO database where the minimum time was 85 days (but the lower quartile 132 days). It is worth noting that because our interest was in the first two doses, we didn’t specify selection based on the brand of the third dose so this may be of a different brand to the first two doses (likely explaining the apparent third AZ doses for many of these cases).

**Table 3.** Distribution of vaccine doses, by brand and DAP.

|  |  |  | N (%) | Time from previous vaccine dose |  |
| --- | --- | --- | --- | --- | --- |
|  |  |  |  | Median (Q1; Q2) | Min; Max |
| ARS | AstraZeneca | Dose 1 | 7 (100) | - | - |
|  |  | Dose 2 | 7 (100) | 84 (76; 84) | 53; 84 |
|  |  | Dose 3 | <5 | - | - |
|  | Janssen | Dose 1 | <5 | - | - |
|  |  | Dose 2 | <5 | - | - |
|  |  | Dose 3 | <5 | - | - |
|  | Moderna | Dose 1 | 42 (100) | - | - |
|  |  | Dose 2 | 34 (80.95) | 41 (28; 42) | 28; 191 |
|  |  | Dose 3 | 12 (28.57) | 183 (150; 201) | 126; 258 |
|  | Pfizer | Dose 1 | 139 (100) | - | - |
|  |  | Dose 2 | 116 (83.45) | 42 (21; 42) | 21; 235 |
|  |  | Dose 3 | 30 (21.58) | 206 (186; 264) | 132; 330 |
| CPRD | AstraZeneca | Dose 1 | 306 (100) | - | - |
|  |  | Dose 2 | 295 (96.41) | 77 (70; 80) | 27; 238 |
|  |  | Dose 3 | 228 (74.51) | 193 (185; 206) | 127; 293 |
|  | Moderna | Dose 1 | 28 (100) | - | - |
|  |  | Dose 2 | 19 (67.86) | 63 (57; 84) | 38; 162 |
|  |  | Dose 3 | 9 (32.14) | 149 (147; 162) | 137; 232 |
|  | Pfizer | Dose 1 | 308 (100) | - | - |
|  |  | Dose 2 | 277 (89.94) | 74 (60; 78) | 19; 322 |
|  |  | Dose 3 | 174 (56.49) | 192 (182; 206) | 117; 307 |
| FISABIO | AstraZeneca | Dose 1 | 16 (100) | - | - |
|  |  | Dose 2 | 15 (93.75) | 82 (72; 84) | 54; 99 |
|  |  | Dose 3 | 7 (43.75) | 164 (158; 172) | 148; 198 |
|  | Janssen | Dose 1 | <5 | - | - |
|  |  | Dose 2 | <5 | - | - |
|  |  | Dose 3 | <5 | - | - |
|  | Moderna | Dose 1 | 45 (100) | - | - |
|  |  | Dose 2 | 40 (88.89) | 28 (28; 28) | 27; 118 |
|  |  | Dose 3 | 17 (37.78) | 190 (132; 199) | 85; 214 |
|  | Pfizer | Dose 1 | 175 (100) | - | - |
|  |  | Dose 2 | 152 (85.98) | 21 (21; 21) | 19; 190 |
|  |  | Dose 3 | 41 (21.34) | 199 (186; 219) | 141; 311 |
| **SIDIAP** | AstraZeneca | Dose 1 | 56 (100) | - | - |
|  |  | Dose 2 | 55 (98.21) | 79 (70; 88) | 54; 110 |
|  |  | Dose 3 | 48 (85.71) | 173 (160; 182) | 126; 250 |
|  | Janssen | Dose 1 | 10 (100) | - | - |
|  |  | Dose 2 | <5 | - | - |
|  |  | Dose 3 | <5 | - | - |
|  | Moderna | Dose 1 | 84 (100) | - | - |
|  |  | Dose 2 | 72 (85.71) | 28 (28; 29) | 28; 168 |
|  |  | Dose 3 | 37 (44.05) | 200 (176; 221) | 90; 318 |
|  | Pfizer | Dose 1 | 254 (100) | - | - |
|  |  | Dose 2 | 230 (90.55) | 21 (21; 22) | 20; 199 |
|  |  | Dose 3 | 146 (57.48) | 206 (191; 226) | 100; 360 |
| **BIFAP-PC** | AstraZeneca | Dose 1 | 18 (100) | - | - |
|  |  | Dose 2 | 16 (88.89) | 78 (73; 84) | 65; 91 |
|  |  | Dose 3 | 13 (72.22) | 163 (154; 171) | 135; 202 |
|  | Janssen | Dose 1 | <5 | - | - |
|  |  | Dose 2 | <5 | - | - |
|  |  | Dose 3 | <5 | - | - |
|  | Moderna | Dose 1 | 59 (100) | - | - |
|  |  | Dose 2 | 46 (77.97) | 28 (28; 30) | 27; 182 |
|  |  | Dose 3 | 12 (20.34) | 189 (171; 230) | 108; 265 |
|  | Pfizer | Dose 1 | 200 (100) | - | - |
|  |  | Dose 2 | 169 (84.5) | 21 (21; 22) | 20; 197 |
|  |  | Dose 3 | 38 (19) | 198 (184; 218) | 132; 326 |

### Graphical Assessment of Model Assumptions

We used exposure-centred interval plots to assess the violation of the event-dependent exposure assumption for the first and second vaccine dose, of each brand, in each country. Not all of these plots could be released due to small numbers and disclosure rules for each data provider. Plots that could be released are provided in the supplementary materials, and an illustrative example from the ARS database is included in Figure 2a-b.

**Figure 2a-b.**
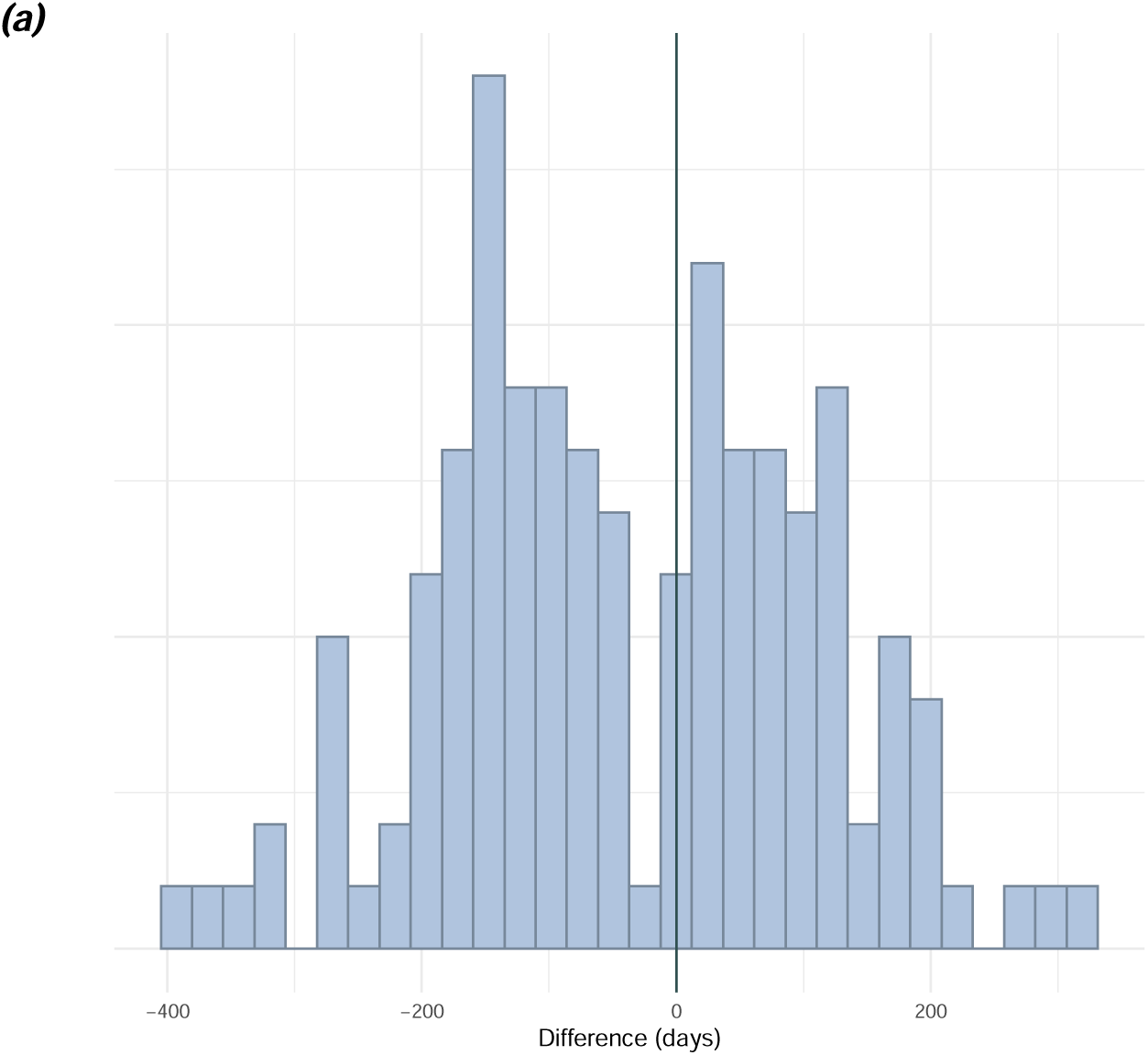
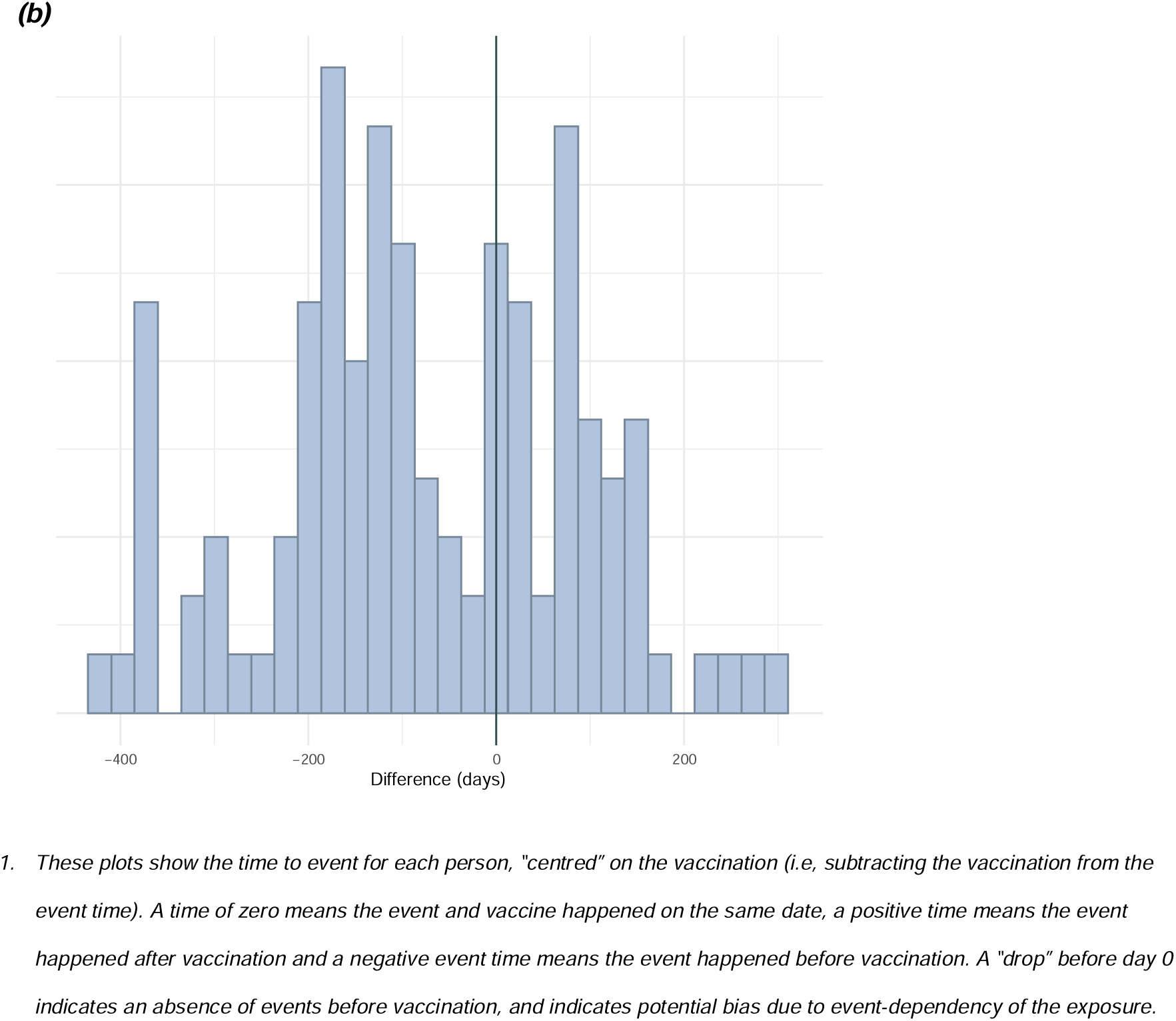
Illustrative exposure-centred intervals plot for Pfizer dose 1 and dose 2 from the ARS database (a-b)^1^.

The key trend observed in these graphical assessments was that there was some evidence of a short but temporary delay of both the first and second doses of Pfizer and Moderna following the occurrence of myocarditis in most countries. This can be seen by a short “drop” in the number of events shortly before both vaccine doses, illustrated by the ARS figures in Figure 1a-b. However, this appeared short-lived and the number of events increased with increasing distance from the vaccine doses. Numbers were generally too low to assess the potential for the delay with the AstraZeneca and Janssen vaccines.

### Regression models comparing the different designs

In unadjusted analyses of the first dose, agreement between designs varied depending on the vaccine brand. The agreement was generally good for AstraZeneca and Pfizer, with no increased risk detected, except for the standard SCCS estimate for Pfizer which indicated a harmful association. For Moderna, an increased rate ratio was found using both a standard and extended SCCS, whereas results from both the pre- and post-vaccination SCRI were consistent with null effects (Figure 3). There was very good agreement between all designs for the unadjusted second dose analyses (Figure 4), demonstrating a harmful association between the second dose of Moderna and Pfizer and myocarditis, and no association for AstraZeneca.

**Figure 3:**
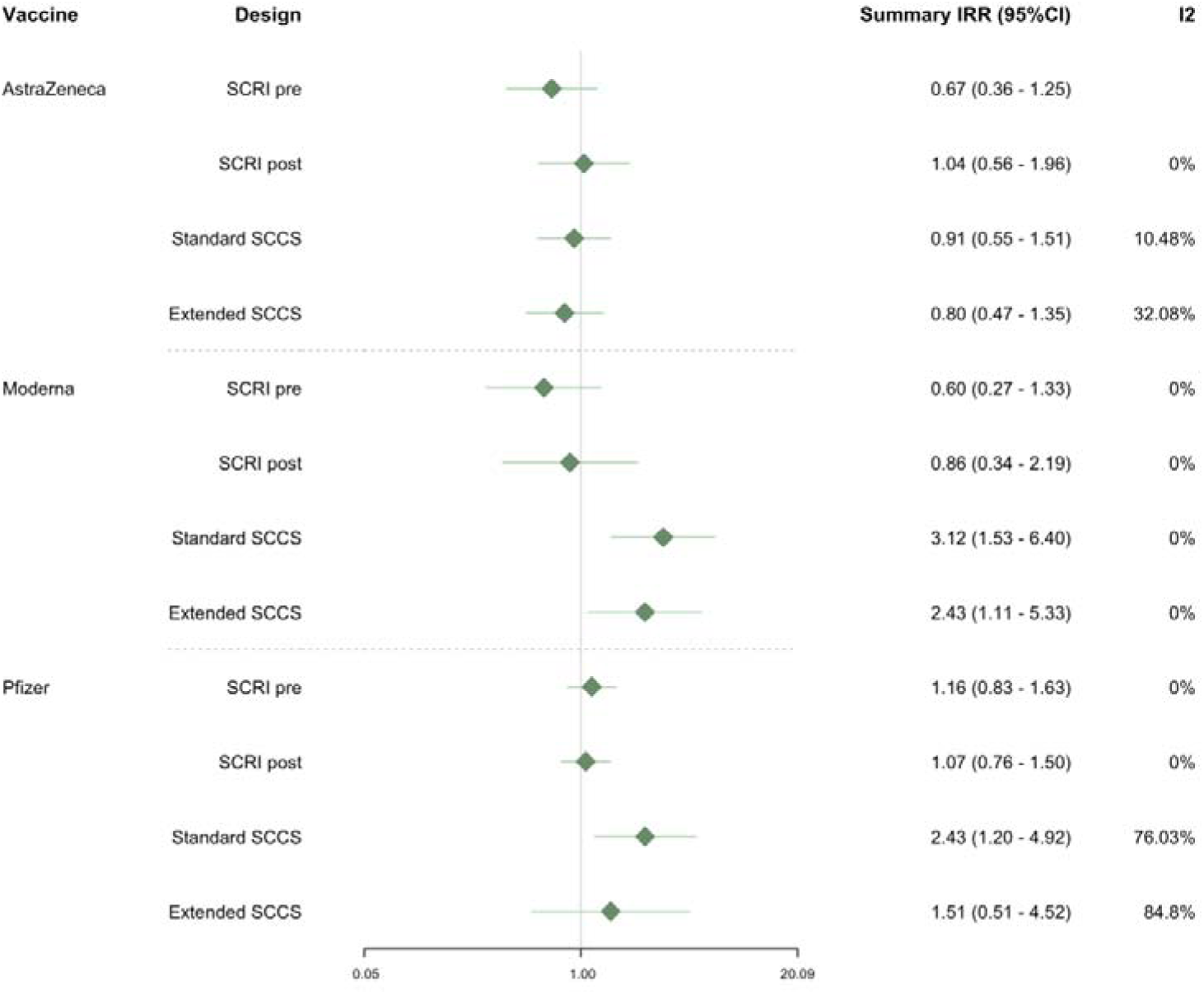
Meta-analysed association between the first dose of each vaccine and myocarditis, unadjusted for calendar time.

**Figure 4:**
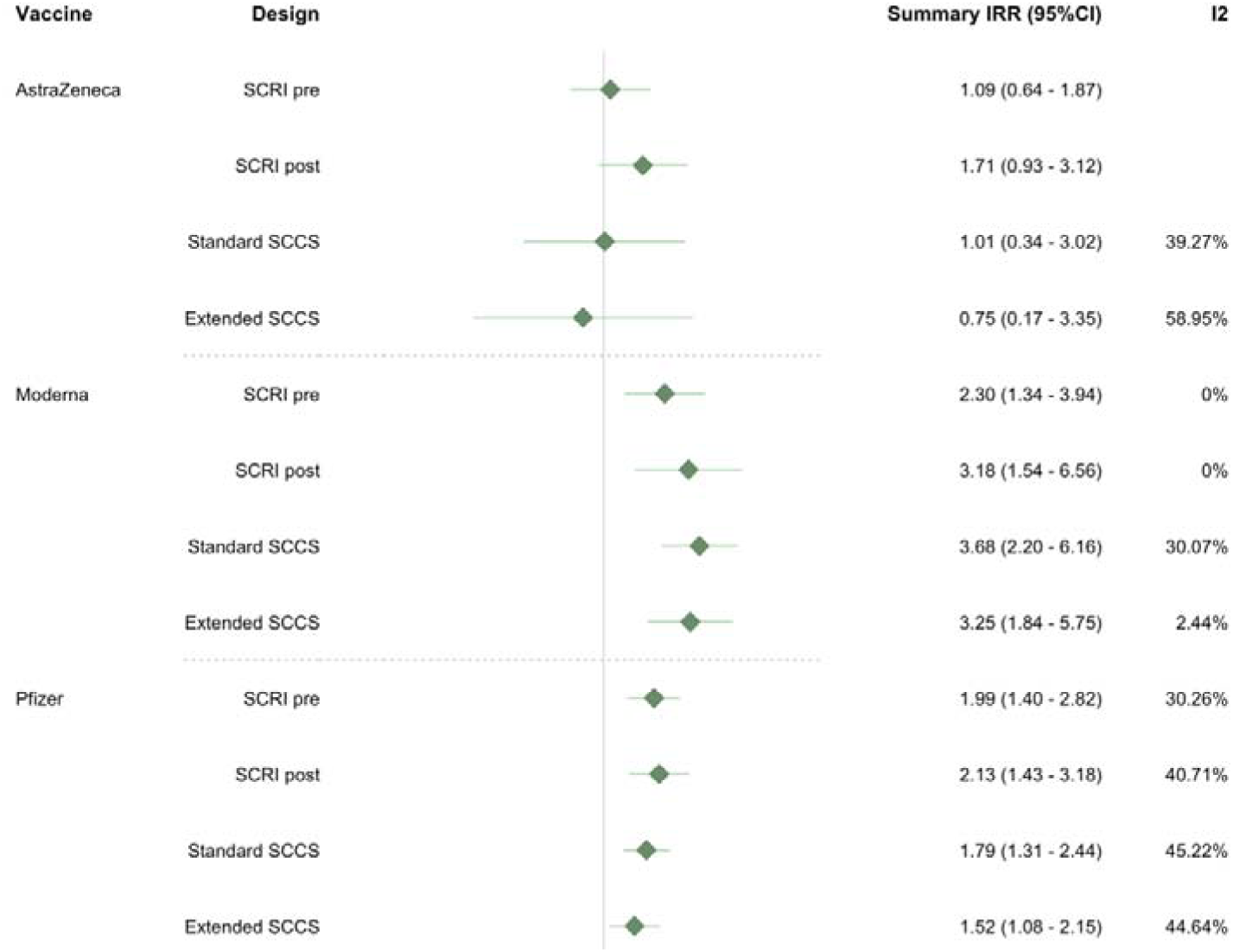
Meta-analysed association between the second dose of each vaccine and myocarditis, unadjusted for calendar time.

Calendar time adjustment was difficult to implement in the SCRIs, and these models suffered from convergence issues and inflated variance which made the designs challenging to compare. For first doses, there was some indication that both the standard and extended SCCS estimated a higher risk of myocarditis after the first dose of Moderna and Pfizer than the pre- and post-vaccination SCRIs, but confidence intervals were very wide (Figure 5).

**Figure 5:**
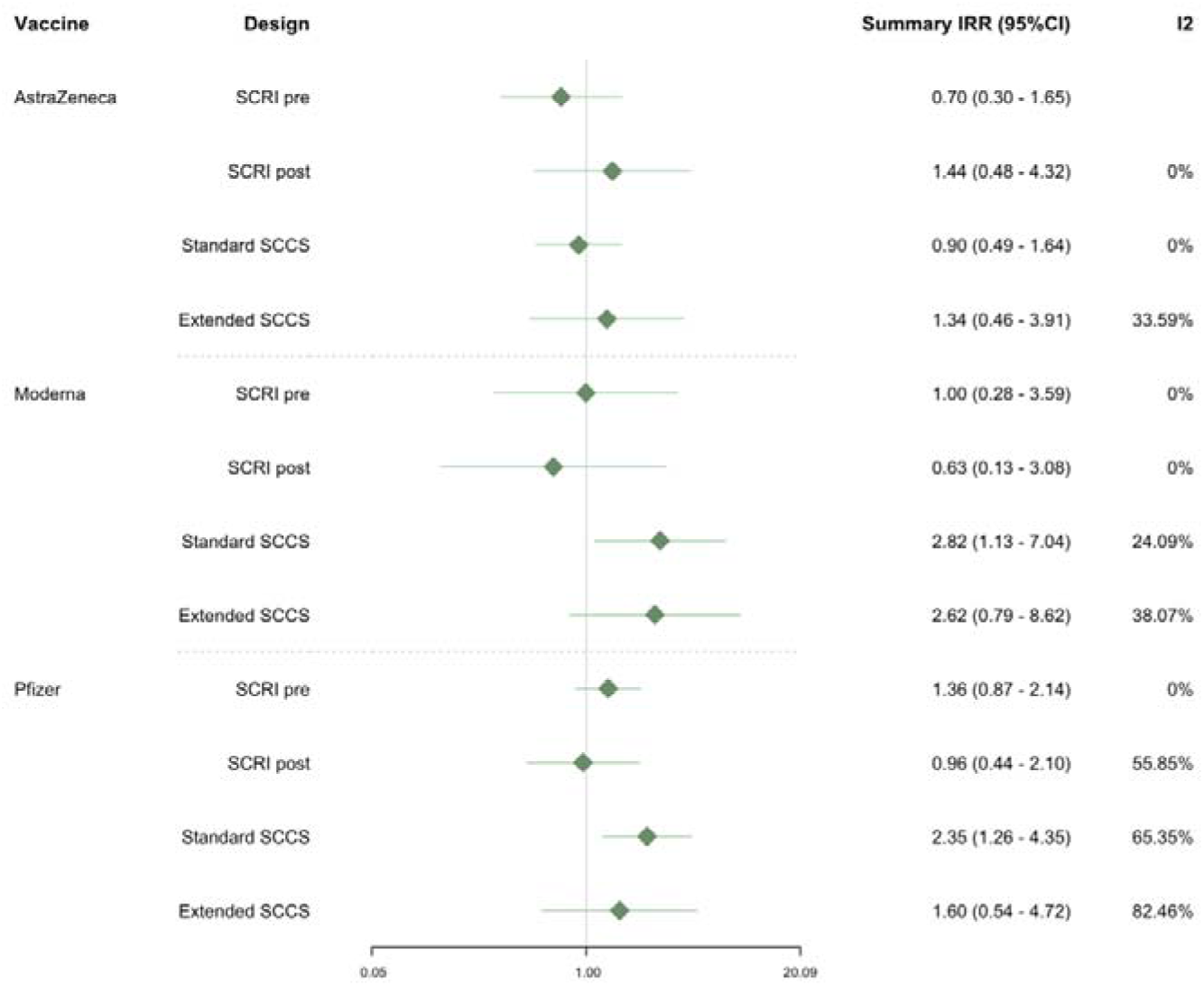
Meta-analysed adjusted association between the first dose of each vaccine and myocarditis, after adjustment for calendar time in 60-day increments.

Agreement between designs was good for AstraZeneca. In adjusted second dose analyses (Figure 6), there appeared to be good agreement between the designs with consistent rate ratios showing no increased risk after the second dose of AstraZeneca, and an increased risk of myocarditis after the second dose of Moderna and the Pfizer vaccines. Country-specific results can be found in the supplementary materials.

**Figure 6:**
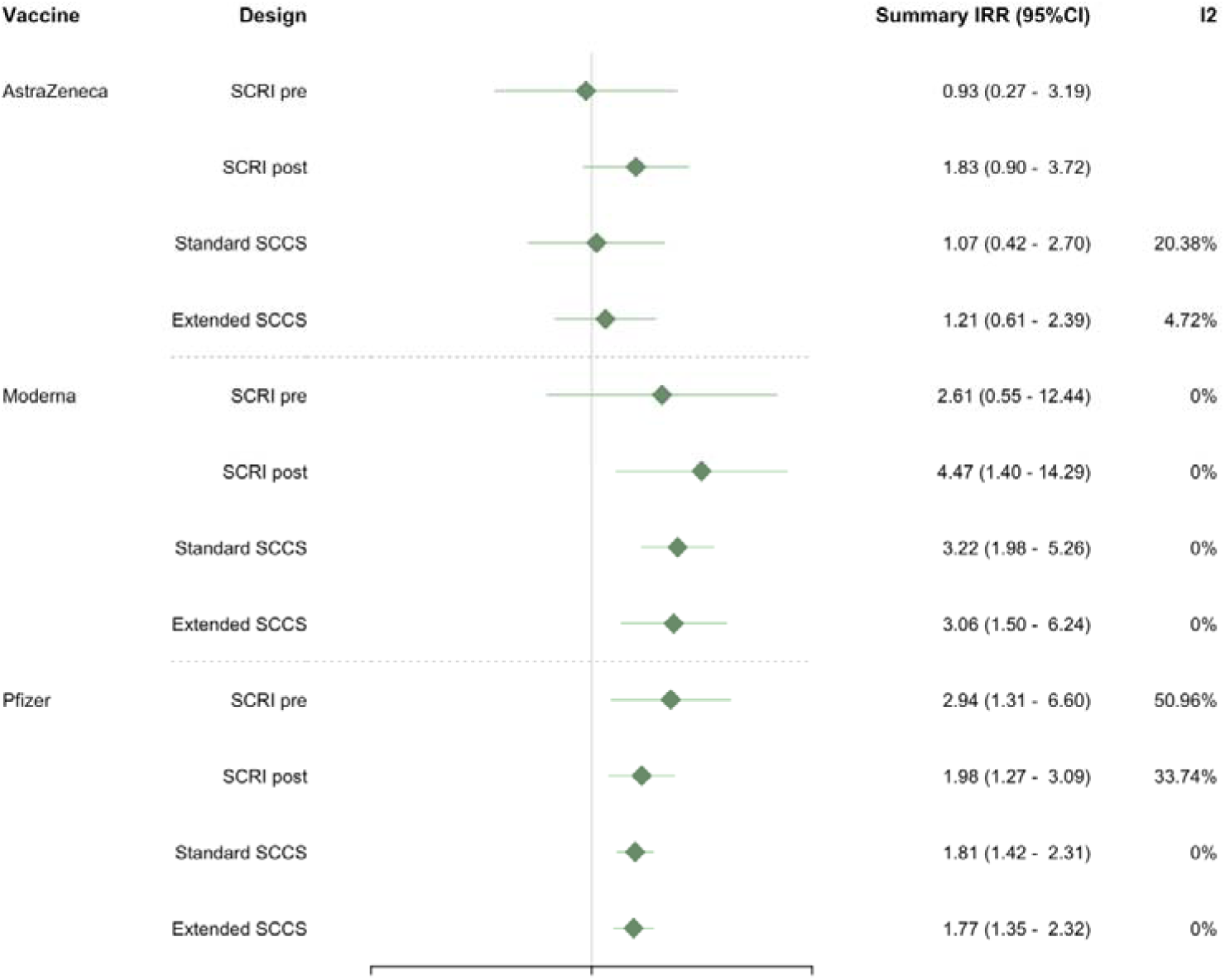
Meta-analysed adjusted association between the second dose of each vaccine and myocarditis, after adjustment for calendar time in 60-day increments.

## Discussion

### Summary

In this study, we compared the ability of four different self-controlled study designs to detect the known association between some COVID-19 vaccines and myocarditis using data from five European countries. All four designs were able to detect the association between the second dose of the Pfizer and Moderna vaccines and an increased risk of myocarditis, although control for calendar time was more challenging in the SCRIs, resulting in wider confidence intervals after covariate adjustment. There was no indication of systematic overestimates of the rate ratio in the pre-SCRI and standard SCCS compared to the post-SCRI or the extended SCCS.

### Comparison to prior literature

Other authors have also compared different study designs for the purpose of detecting vaccine safety effects, most recently Schuemie et al in an evaluation of 25 different study designs in four different US health insurance databases (2). The evaluation included a comparison between a standard SCCS, post-vaccine SCCS, pre-vaccine SCRI, and post-vaccine SCRI. Performance was compared in terms of the ability of each method to detect associations between six different vaccines and three simulated positive controls; and their ability to not detect associations for 93 l negative controls. This resulted in over a million effect size estimates, on which performance metrics such as type I and type II errors were computed. The authors concluded that the self-controlled methods generally performed better than historical cohort and case-control methods given the performance metrics calculated in the paper, although the SCCS and SCRI were not explicitly compared (2).

From the data presented performance appears relatively similar, although multiple vaccine doses were considered as separate individuals in this analysis and therefore potential violations of the self-controlled study assumptions could not be addressed.

Focusing specifically on the choice of design in the context of violations of the event-dependent exposure assumption, Hua et al evaluated the performance of the standard SCCS, a post-vaccination SCCS and the extended SCCS in a simulation study of two vaccinations and a rare adverse event (8). They found that for a single vaccine dose and no seasonal confounders, the post-vaccination and extended SCCS were both unbiased whereas use of the standard SCCS resulted in an upward bias. However, in the multidose setting, only the extended SCCS was able to recover unbiased estimates for both vaccine doses (8).

### Interpretation

Taken together, our results extend findings from previous comparisons of study designs for determining vaccine safety by providing a detailed comparison of different SCCS and SCRI design options in a clinical setting where we expected exposures to be event-dependent.

Our key finding is that there was generally good agreement between these designs, implying that the extent of violations of this assumption might be limited in our setting. This was confirmed through a graphical investigation, which suggested that any violation of the assumption of event-dependent exposures was likely to be short-lived. Our results are therefore in agreement with those from Schuemie and colleagues, which showed similar performance of both SCRI and SCCS designs (2)

An interesting and unexpected finding was that the Moderna and Pfizer first dose rate ratios were somewhat higher using both the standard and extended SCCS, with both of the SCRI finding results closer to the null. This was particularly marked before the adjustment for calendar time. Although findings should be interpreted cautiously as confidence intervals overlapped between the different study designs, the SCCS may be more sensitive to calendar time trends than the SCRI, as it includes a significantly longer follow-up period than the SCRIs. An increase in the rate of myocarditis over time, for example, due to increasing COVID-19 prevalence, might have been expected to produce the observed results. Nevertheless, differences in study designs were not consistent between doses, and we cannot rule out that some of the variation may be due to random error.

Another difference worth commenting on is that although both the standard and extended SCCS included a greater number of events, we found wider confidence intervals for AstraZeneca adjusted first dose estimates using these designs than using either the pre- or post-vaccine SCRIs. This appeared to be due to the fact that some SCCS, but not SCRI, ran with a very low number of events in databases where the AstraZeneca vaccines were not widely used. These estimates therefore contributed to the meta-analyses for the SCCS, but not for the SCRI. Comparing estimates in the CPRD database only showed narrower confidence intervals for the SCCS than the SCRI for these comparisons, as expected.In general, power in self-controlled studies relies not only on the number of events but on the ratio of risk to control time (13), although a simulation study specifically comparing the power of an SCCS vs the SCRI found that the SCRI generally had less power than the SCCS (14).

When choosing a certain study design, there are also important pragmatic considerations to take into account. For example, the post-vaccination SCRI required the accumulation of sufficient time after the second vaccine dose, and for urgent questions, it may not be feasible to wait for follow-up time to accumulate. The SCCS may offer practical advantages compared to SCRI designs when attempting to adjust for covariates. We struggled to control for calendar time in the country-specific SCRI due to a low number of events and a relatively short follow-up period (<120 days), which resulted in collinearity between the proposed calendar time variables and the risk intervals and in turn very wide confidence intervals in the SCRI, but not the SCCS. There are alternative options for adjusting for calendar time in an SCRI (15), and these options may be useful where time-varying confounding by calendar time is a particular concern. More generally, time-varying confounding may be less problematic in an SCRI as the observation periods are shorter than in the SCCS.

### Limitations

Within each country, there was often a small number of cases available. This was reflected in convergence problems and increased variance in the post-vaccine SCRI for some vaccines and countries. We also found that currently available software for fitting extensions of the SCCS did not always notify the user of convergence issues, and had limited options for covariate adjustment. An important avenue for future work may be to improve software for fitting SCCS extensions. Our experience also highlights that there may be pragmatic reasons for using a simpler, design-based solution such as a truncated SCCS or post-vaccine SCRI to tackle the issue of event-dependent exposures. It’s also important to note that we only compared four possible design options that we considered particularly relevant for our case study. There are of course several other potential study designs that could be used to address the study question, including case-crossover designs, or between-person study designs such as cohort or case-control studies. The most reliable inferences can likely be made by triangulating findings from multiple different study designs, subject to different underlying assumptions (16). Finally, it should be noted that our results here were generated for methodological purposes and we therefore did not stratify our analyses by age, or study heterologous dosing. This means that we caution against a clinical interpretation of these results, particularly as strong effect modification by age has been shown for this safety signal (11).

## Conclusions

In the context of the known association between COVID-19 vaccines and myocarditis, we have demonstrated that two forms of SCRI and two forms of SCCS led to largely comparable results. This is likely because there was a limited violation of the assumption of event-dependent exposures in all contributing data sources and relatively limited variation in the recording of the outcome over time, and, as a result, the four designs appear equally valid for evaluating this specific clinical question. Pragmatically, the SCCS may offer some advantages compared to the SCRI when there are important time-varying confounders and a low number of cases, as the models may be easier to fit. In cases where time-varying confounding is less of a concern than event-dependency of the exposure, our evaluation suggests that using a post-vaccination SCRI – specifying a control window either after the final dose or in between two doses separated by some minimum distance – may be a useful option. A simulation study of these designs under the presence of multiple different biases, and considering more complex vaccination schedules, would be a valuable area for future research.

## Data Availability

The datasets used in these study contain confidential patient information and cannot be shared. Code for preparing the data is available here: https://github.com/VAC4EU/CVM. The code for running these specific analyses will be made available on Github (note: link to be made public on paper acceptance; peer-reviewers or interested parties can be given earlier access on request).

## Conflicts of Interest

AS is employeed by LSHTM on a fellowship sponsored by GSK. ID owns shares in and reports research grants from GSK, and research grants from AstraZeneca, both unrelated to the current work. FR is an employee of TEAMIT Institute, a research management organisation that participates in financially supported studies for the European Medicines Agency and related healthcare authorities, pharmaceutical companies, and the European Union. FV, MPM, CAB and ES are salaried employees at Fundació Institut Universitari per a la recerca a l’Atenció Primària de Salut Jordi Gol i Gurina (IDIAPJGol), which receives institutional research funding from public and private partners, pharmaceutical companies and regulatory agencies, administered by IDIAPJGol. MS is head of a department that conducts studies for the European Medicines Agency, the European Commission and medicine manufacturers, all according to the ENCePP code of conduct. MS does not hold personal financial relations with the companies. Carlos E. Durán (CED) is salaried employee by University Medical Center Utrecht, the Netherlands, which receives institutional research funding from pharmaceutical companies and regulatory agencies. CED is involved only in research projects funded by regulatory authorities. RG and DM are employees of ARS Tuscany, which reports funding from the Innovative Medicines Initiative, RTI, PHARMO, University of Southern Denmark, University of Utrecht, Eli Lilly, Pfizer, Novartis, AstraZeneca, Galapagos, and LeoPharma, for studies unrelated to the current work, and conducted in compliance with the ENCePP code of conduct.

## Ethics

This work was conducted as part of a larger programme of work, and this specific methodological sub-study was approved by the LSHTM ethics committee (ref 28222). The larger programme of work had ethics approval from each contributing DAP, as shown below. The protocol was pre-registered on ENCEPP (EUPAS42467).

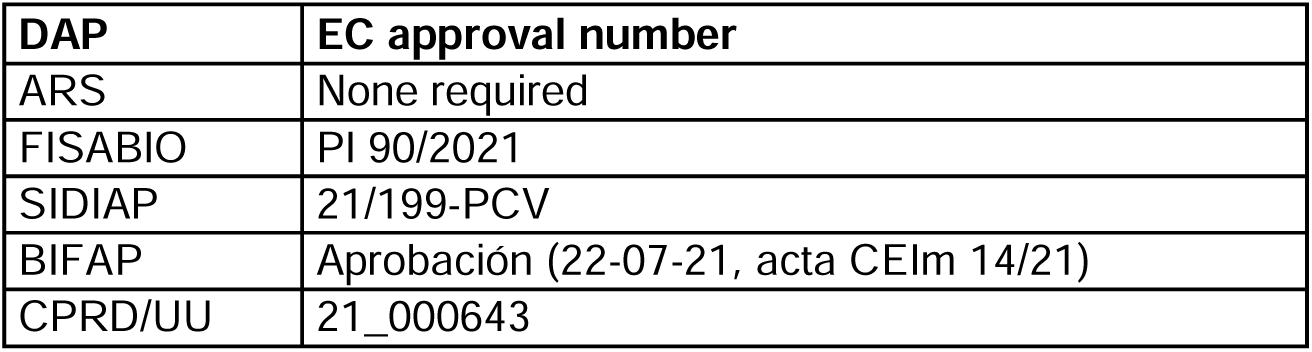

| DAP | EC approval number |
| --- | --- |
| ARS | None required |
| FISABIO | PI 90/2021 |
| SIDIAP | 21/199-PCV |
| BIFAP | Aprobación (22-07-21, acta CEIm 14/21) |
| CPRD/UU | 21_000643 |

## Footnotes

1 These plots are “centred” on vaccination, that is, the time of vaccination is subtracted from each event time and the histogram therefore displays the event times relative to each vaccination date. Time zero is the time of each vaccination, a positive time represents an event occurring after vaccination, and a negative time an event occurring before vaccination.

## Notes

### Competing Interest Statement

Conflicts of Interest
AS is employed by LSHTM on a fellowship sponsored by GSK. ID owns shares in and reports research grants from GSK, and research grants from AstraZeneca, both unrelated to the current work. FR is an employee of TEAMIT Institute, a research management organisation that participates in financially supported studies for the European Medicines Agency and related healthcare authorities, pharmaceutical companies, and the European Union. Felipe Villalobos, Meritxell Palleja-Millan, Carlo Alberto Bissacco and Elena Segundo are salaried employees at Fundacio Institut Universitari per a la recerca a l'Atencio Primaria de Salut Jordi Gol i Gurina (IDIAPJGol), which receives institutional research funding from public and private partners, pharmaceutical companies and regulatory agencies, administered by IDIAPJGol. MS is head of a department that conducts studies for the European Medicines Agency, the European Commission and medicine manufacturers, all according to the ENCePP code of conduct. MS does not hold personal financial relations with the companies. Carlos E. Duran (CED) is salaried employee by University Medical Center Utrecht, the Netherlands, which receives institutional research funding from pharmaceutical companies and regulatory agencies. CED is involved only in research projects funded by regulatory authorities. RG and DM are employees of ARS Tuscany, which reports funding from the Innovative Medicines Initiative, RTI, PHARMO, University of Southern Denmark, University of Utrecht, Eli Lilly, Pfizer, Novartis, AstraZeneca, Galapagos, and LeoPharma, for studies unrelated to the current work, and conducted in compliance with the ENCePP code of conduct.

### Author Declarations

This work was conducted as part of a larger programme of work, and this specific methodological sub-study was approved by the LSHTM ethics committee (ref 28222). The larger programme of work had ethics approval from each contributing DAP, as shown below. The protocol was pre-registered on ENCEPP (EUPAS42467). DAPEC approval number ARSNone required FISABIOPI 90/2021 SIDIAP21/199-PCV BIFAPAprobacion (22-07-21, acta CEIm 14/21) CPRD/UU21_000643

