## supplemental materials for "A comparison of four self-controlled study designs in an analysis of COVID-19 vaccines and Myocarditis using Five European Databases"

**APPENDIX**

**Table 1. Results for each method in the ARS**

|  |  | **Unadjusted** | | |  | **Adjusted** | | | |
| --- | --- | --- | --- | --- | --- | --- | --- | --- | --- |
|  |  | **Pre-SCRI** | **Post-SCRI** | **SCCS** | **Extended SCCS** | **Pre-SCRI** | **Post-SCRI** | **SCCS** | **Extended SCCS** |
|  |  | RR (95%CI) | RR (95%CI) | RR (95%CI) | RR (95%CI) | RR (95%CI) | RR (95%CI) | RR (95%CI) | RR (95%CI) |
| AstraZeneca | Dose 1 | <5 cases, not run | <5 cases, not run | did not converge | did not converge | <5 cases, not run | <5 cases, not run | did not converge | did not converge |
|  | Dose 2 | <5 cases, not run | <5 cases, not run | did not converge | did not converge | <5 cases, not run | <5 cases, not run | did not converge | did not converge |
| Janssen | Dose 1 | <5 cases, not run | <5 cases, not run | <5 cases, not run | <5 cases, not run | <5 cases, not run | <5 cases, not run | <5 cases, not run | <5 cases, not run |
|  | Dose 2 | <5 cases, not run | <5 cases, not run | <5 cases, not run | <5 cases, not run | <5 cases, not run | <5 cases, not run | <5 cases, not run | <5 cases, not run |
| Moderna | Dose 1 | 0.31 (0.04-2.48) | did not converge | 1.08 (0.14-8.08) | did not converge | did not converge | did not converge | 0.83 (0.11-6.49) | did not converge |
|  | Dose 2 | 3.14 (1.07-9.2) | did not converge | 4.57 (2.05-10.18) | 5.09 (2.2-11.78) | did not converge | did not converge | 3.6 (1.44-8.97) | did not converge |
| Pfizer | Dose 1 | 1.81 (0.85-3.87) | 1.5 (0.69-3.23) | 3.54 (1.86-6.73) | 2.92 (1.51-5.66) | 3.57 (1.05-12.09) | 4.09 (1.18-14.15) | 3 (1.51-5.95) | 1.85 (0.91-3.76) |
|  | Dose 2 | 2.74 (1.25-6.02) | 2.77 (1.25-6.15) | 2.04 (1.14-3.65) | 2.04 (1-4.14) | 7.24 (1.57-33.37) | 5.24 (1.88-14.61) | 1.89 (1.03-3.46) | 1.65 (0.83-3.3) |

**Table 2. Results for each method in SIDIAP**

|  |  | **Unadjusted** | | |  | **Adjusted** | | | |
| --- | --- | --- | --- | --- | --- | --- | --- | --- | --- |
|  |  | **Pre-SCRI** | **Post-SCRI** | **SCCS** | **Extended SCCS** | **Pre-SCRI** | **Post-SCRI** | **SCCS** | **Extended SCCS** |
|  |  | RR (95%CI) | RR (95%CI) | RR (95%CI) | RR (95%CI) | RR (95%CI) | RR (95%CI) | RR (95%CI) | RR (95%CI) |
| AstraZeneca | Dose 1 | <5 cases, not run | <5 cases, not run | 0.36 (0.05-2.59) | 0.25 (0.03-1.82) | <5 cases, not run | <5 cases, not run | 0.86 (0.07-9.92) | 1.67 (0.15-18.88) |
|  | Dose 2 | <5 cases, not run | <5 cases, not run | 0.36 (0.05-2.62) | 0.25 (0.03-1.81) | <5 cases, not run | <5 cases, not run | 0.34 (0.04-3.3) | 0.39 (0.04-3.81) |
| Janssen | Dose 1 | <5 cases, not run | <5 cases, not run | <5 cases, not run | <5 cases, not run | <5 cases, not run | <5 cases, not run | <5 cases, not run | <5 cases, not run |
|  | Dose 2 | <5 cases, not run | <5 cases, not run | <5 cases, not run | <5 cases, not run | <5 cases, not run | <5 cases, not run | <5 cases, not run | <5 cases, not run |
| Moderna | Dose 1 | 0.43 (0.09-1.96) | 0.84 (0.16-4.34) | 2.98 (0.68-13.04) | 2.8 (0.67-11.68) | 1.82 (0.18-17.91) | 0.45 (0.04-5.61) | 3.46 (0.76-15.79) | 4.16 (0.88-19.77) |
|  | Dose 2 | 1.68 (0.59-4.82) | 4.44 (1.16-17.05) | 1.81 (0.78-4.19) | 1.86 (0.75-4.6) | 6.62 (0.41-105.54) | 4.63 (0.72-29.71) | 2.08 (0.83-5.21) | 2.97 (1.11-7.99) |
| Pfizer | Dose 1 | 1.25 (0.61-2.58) | 1.15 (0.55-2.37) | 4.86 (2-11.81) | 3.32 (1.25-8.8) | 1.15 (0.4-3.33) | 1.16 (0.39-3.48) | 5.16 (2.07-12.86) | 5.47 (1.98-15.06) |
|  | Dose 2 | 1.07 (0.5-2.27) | 1.04 (0.49-2.21) | 0.96 (0.51-1.81) | 0.75 (0.39-1.43) | 0.84 (0.23-3.09) | 0.99 (0.38-2.58) | 1.06 (0.54-2.07) | 1.26 (0.62-2.55) |

**Table 3. Results for each method in FISABIO**

|  |  | **Unadjusted** | | |  | **Adjusted** | | | |
| --- | --- | --- | --- | --- | --- | --- | --- | --- | --- |
|  |  | **Pre-SCRI** | **Post-SCRI** | **SCCS** | **Extended SCCS** | **Pre-SCRI** | **Post-SCRI** | **SCCS** | **Extended SCCS** |
|  |  | RR (95%CI) | RR (95%CI) | RR (95%CI) | RR (95%CI) | RR (95%CI) | RR (95%CI) | RR (95%CI) | RR (95%CI) |
| AstraZeneca | Dose 1 | <5 cases, not run | <5 cases, not run | did not converge | did not converge | <5 cases, not run | <5 cases, not run | did not converge | did not converge |
|  | Dose 2 | <5 cases, not run | <5 cases, not run | did not converge | did not converge | <5 cases, not run | <5 cases, not run | did not converge | did not converge |
| Janssen | Dose 1 | <5 cases, not run | <5 cases, not run | <5 cases, not run | <5 cases, not run | <5 cases, not run | <5 cases, not run | <5 cases, not run | <5 cases, not run |
|  | Dose 2 | <5 cases, not run | <5 cases, not run | <5 cases, not run | <5 cases, not run | <5 cases, not run | <5 cases, not run | <5 cases, not run | <5 cases, not run |
| Moderna | Dose 1 | 0.37 (0.04-3.04) | 0.27 (0.03-2.2) | did not converge | did not converge | did not converge | did not converge | did not converge | did not converge |
|  | Dose 2 | 1.76 (0.54-5.75) | 1.71 (0.53-5.51) | did not converge | did not converge | did not converge | did not converge | did not converge | did not converge |
| Pfizer | Dose 1 | 0.53 (0.18-1.53) | 0.56 (0.17-1.85) | 0.55 (0.07-4.04) | 0.69 (0.09-5.21) | 1.25 (0.31-4.99) | 0.34 (0.07-1.64) | 0.18 (0.05-0.63) | 0.23 (0.07-0.77) |
|  | Dose 2 | 1.61 (0.81-3.21) | 2.09 (0.99-4.45) | 1.6 (0.92-2.78) | 1.96 (1.08-3.56) | 5.51 (1.3-23.31) | 1.47 (0.51-4.29) | 1.62 (0.85-3.09) | 2.12 (1.09-4.15) |

**Table 4. Results for each method in CPRD**

|  |  | **Unadjusted** | | |  | **Adjusted** | | | |
| --- | --- | --- | --- | --- | --- | --- | --- | --- | --- |
|  |  | **Pre-SCRI** | **Post-SCRI** | **SCCS** | **Extended SCCS** | **Pre-SCRI** | **Post-SCRI** | **SCCS** | **Extended SCCS** |
|  |  | RR (95%CI) | RR (95%CI) | RR (95%CI) | RR (95%CI) | RR (95%CI) | RR (95%CI) | RR (95%CI) | RR (95%CI) |
| AstraZeneca | Dose 1 | 0.67 (0.36-1.25) | 1.04 (0.53-2.05) | 0.87 (0.49-1.52) | 0.77 (0.43-1.38) | 0.7 (0.3-1.65) | 1.45 (0.46-4.58) | 0.81 (0.42-1.54) | 0.79 (0.4-1.54) |
|  | Dose 2 | 1.09 (0.64-1.87) | 1.71 (0.93-3.12) | 1.37 (0.87-2.17) | 1.27 (0.77-2.11) | 0.93 (0.27-3.19) | 1.83 (0.9-3.72) | 1.3 (0.74-2.27) | 1.35 (0.73-2.49) |
| Janssen | Dose 1 | <5 cases, not run | <5 cases, not run | <5 cases, not run | <5 cases, not run | <5 cases, not run | <5 cases, not run | <5 cases, not run | <5 cases, not run |
|  | Dose 2 | <5 cases, not run | <5 cases, not run | <5 cases, not run | <5 cases, not run | <5 cases, not run | <5 cases, not run | <5 cases, not run | <5 cases, not run |
| Moderna | Dose 1 | did not converge | did not converge | 5.6 (2.01-15.61) | 3.45 (1.05-11.31) | did not converge | did not converge | 7.63 (1.89-30.75) | 5.75 (1.01-32.74) |
|  | Dose 2 | did not converge | did not converge | 7.2 (2.19-23.66) | 2.12 (0.42-10.87) | did not converge | did not converge | 4.51 (1.17-17.43) | 1.92 (0.33-11.3) |
| Pfizer | Dose 1 | 1.15 (0.64-2.06) | 0.98 (0.55-1.76) | 1.08 (0.65-1.8) | 0.84 (0.49-1.44) | 1.05 (0.52-2.13) | 0.98 (0.44-2.17) | 1.17 (0.69-1.99) | 0.91 (0.51-1.61) |
|  | Dose 2 | 1.92 (1.13-3.28) | 1.92 (1.12-3.29) | 1.78 (1.18-2.7) | 1.45 (0.93-2.27) | 1.59 (0.59-4.29) | 1.81 (1-3.28) | 1.87 (1.2-2.91) | 1.59 (0.98-2.59) |

**Table 5. Results for each method in BIFAP**

|  |  | **Unadjusted** | | |  | **Adjusted** | | | |
| --- | --- | --- | --- | --- | --- | --- | --- | --- | --- |
|  |  | **Pre-SCRI** | **Post-SCRI** | **SCCS** | **Extended SCCS** | **Pre-SCRI** | **Post-SCRI** | **SCCS** | **Extended SCCS** |
|  |  | RR (95%CI) | RR (95%CI) | RR (95%CI) | RR (95%CI) | RR (95%CI) | RR (95%CI) | RR (95%CI) | RR (95%CI) |
| AstraZeneca | Dose 1 | did not converge | 1.07 (0.2-5.85) | 2.2 (0.5-9.78) | 2.29 (0.45-11.54) | did not converge | 1.39 (0.04-51.96) | 2.85 (0.36-22.58) | 3.83 (0.69-21.21) |
|  | Dose 2 | did not converge | <5 cases, not run | <5 cases, not run | <5 cases, not run | did not converge | <5 cases, not run | <5 cases, not run | <5 cases, not run |
| Janssen | Dose 1 | <5 cases, not run | <5 cases, not run | <5 cases, not run | <5 cases, not run | <5 cases, not run | <5 cases, not run | <5 cases, not run | <5 cases, not run |
|  | Dose 2 | <5 cases, not run | <5 cases, not run | <5 cases, not run | <5 cases, not run | <5 cases, not run | <5 cases, not run | <5 cases, not run | <5 cases, not run |
| Moderna | Dose 1 | 1.08 (0.32-3.58) | 1.43 (0.36-5.58) | 1.89 (0.43-8.26) | 1.13 (0.24-5.31) | 0.75 (0.16-3.56) | 0.79 (0.1-6.06) | 1.54 (0.32-7.34) | 0.81 (0.17-3.95) |
|  | Dose 2 | 2.83 (1.02-7.84) | 4.89 (1.37-17.39) | 3.99 (1.83-8.68) | 4.32 (1.46-12.8) | 1.69 (0.25-11.2) | 4.36 (0.98-19.39) | 3.8 (1.54-9.36) | 4.07 (1.14-14.48) |
| Pfizer | Dose 1 | 1.03 (0.45-2.34) | 1.07 (0.47-2.46) | 4.12 (1.8-9.45) | 4.61 (1.6-13.22) | 1.46 (0.49-4.36) | 0.41 (0.13-1.32) | 3.22 (1.38-7.56) | 4.87 (1.55-15.28) |
|  | Dose 2 | 3.18 (1.69-5.99) | 3.74 (1.91-7.33) | 2.73 (1.73-4.29) | 2.14 (1.32-3.48) | 5.55 (1.72-17.92) | 2.4 (1.02-5.63) | 2.15 (1.3-3.57) | 2.31 (1.33-3.99) |

**Exposure-Centred Interval Plots**

These plots show the time to event for each person, “centred” on the vaccination (i.e, subtracting the vaccination from the event time). A time of zero means the event and vaccine happened on the same date, a positive time means the event happened after vaccination and a negative event time means the event happened before vaccination. A “drop” before day 0 indicates an absence of events before vaccination, and indicates potential bias due to event-dependency of the exposure.

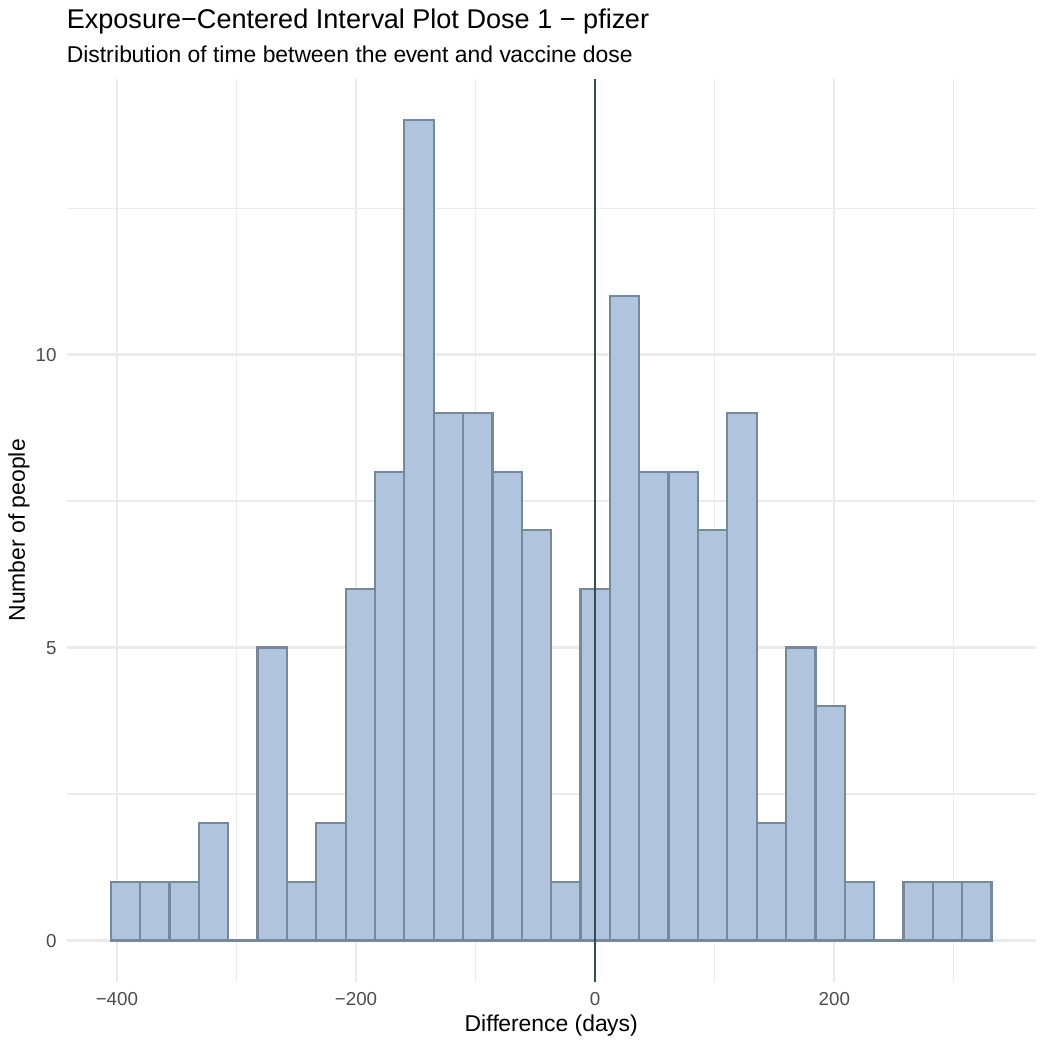
**Figure** **1. ARS Pfizer Dose 1 and 2**

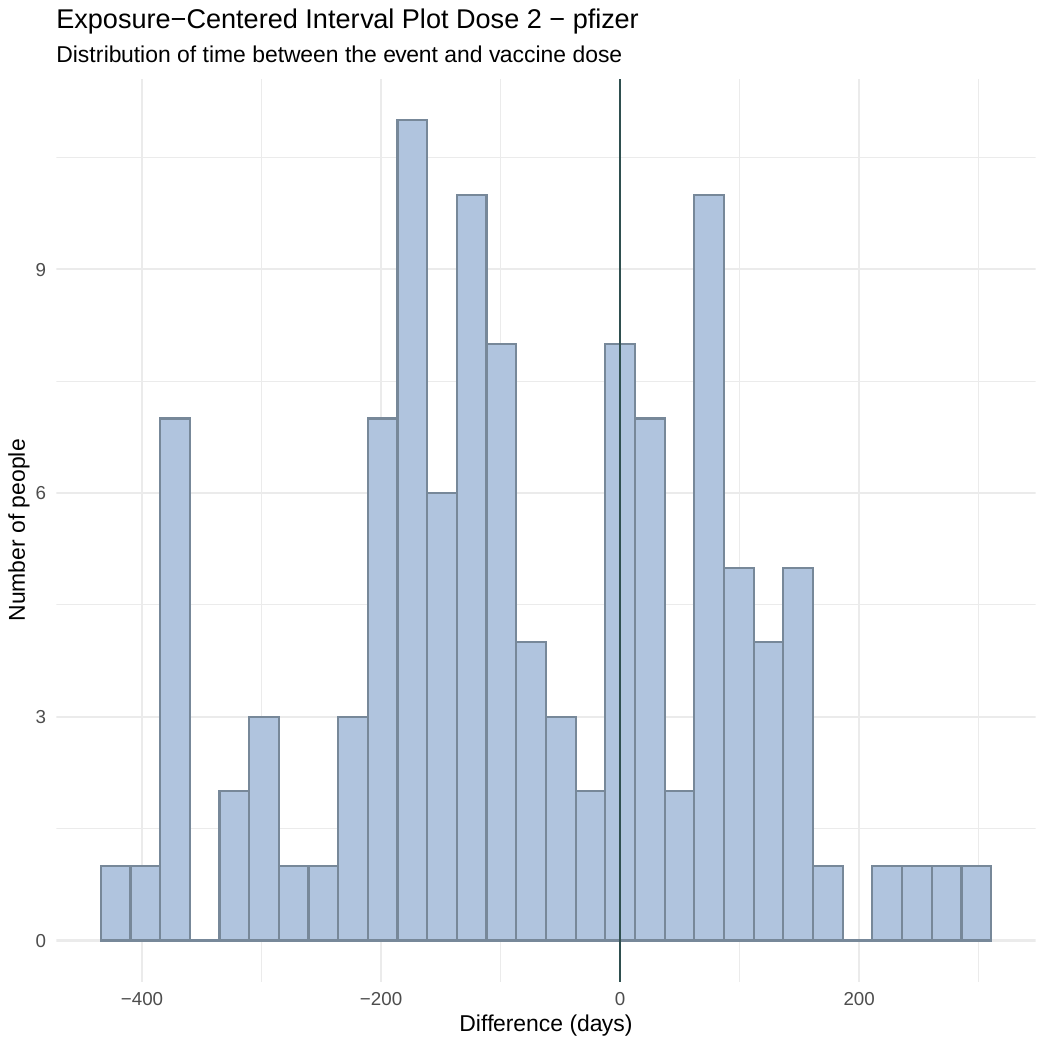

***Figure 2. FISABIO Pfizer Dose 1 and 2***

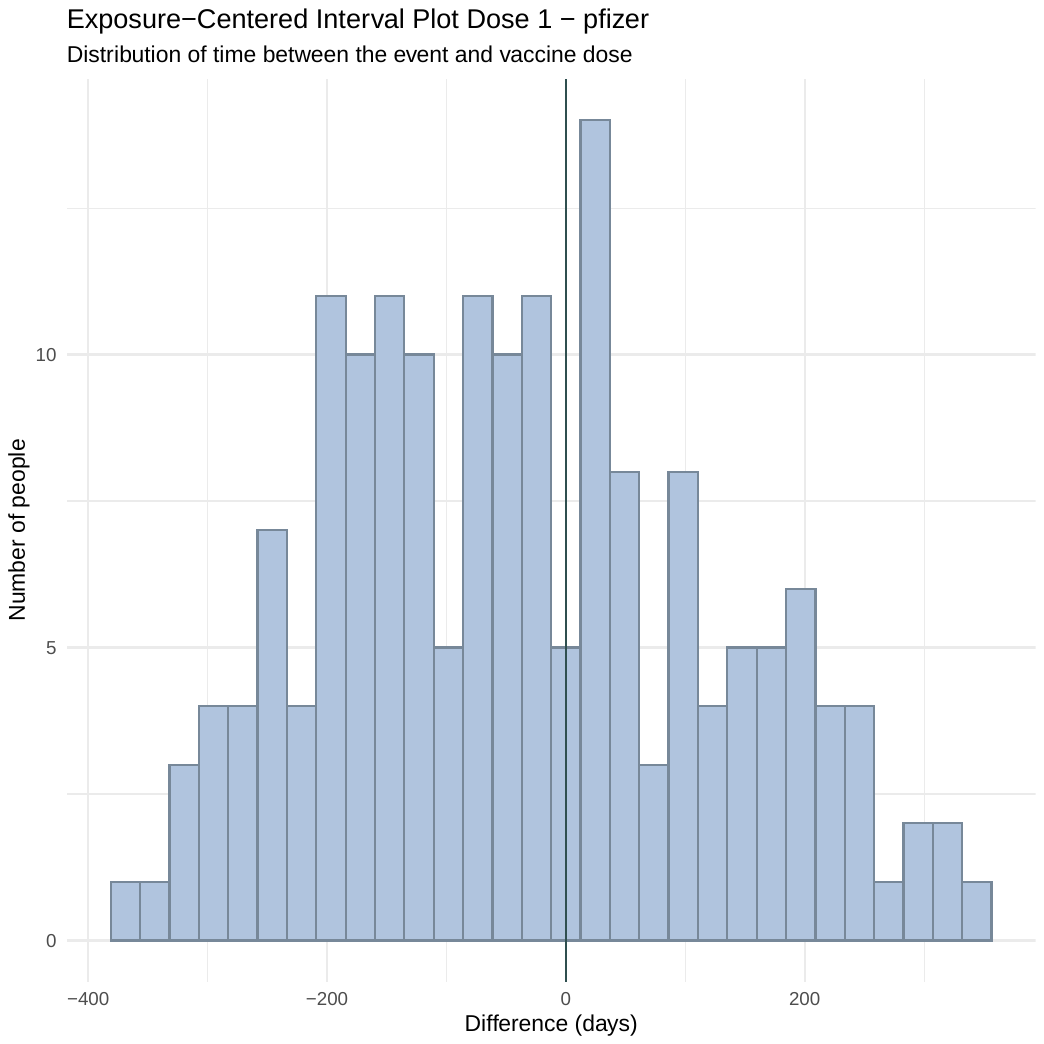

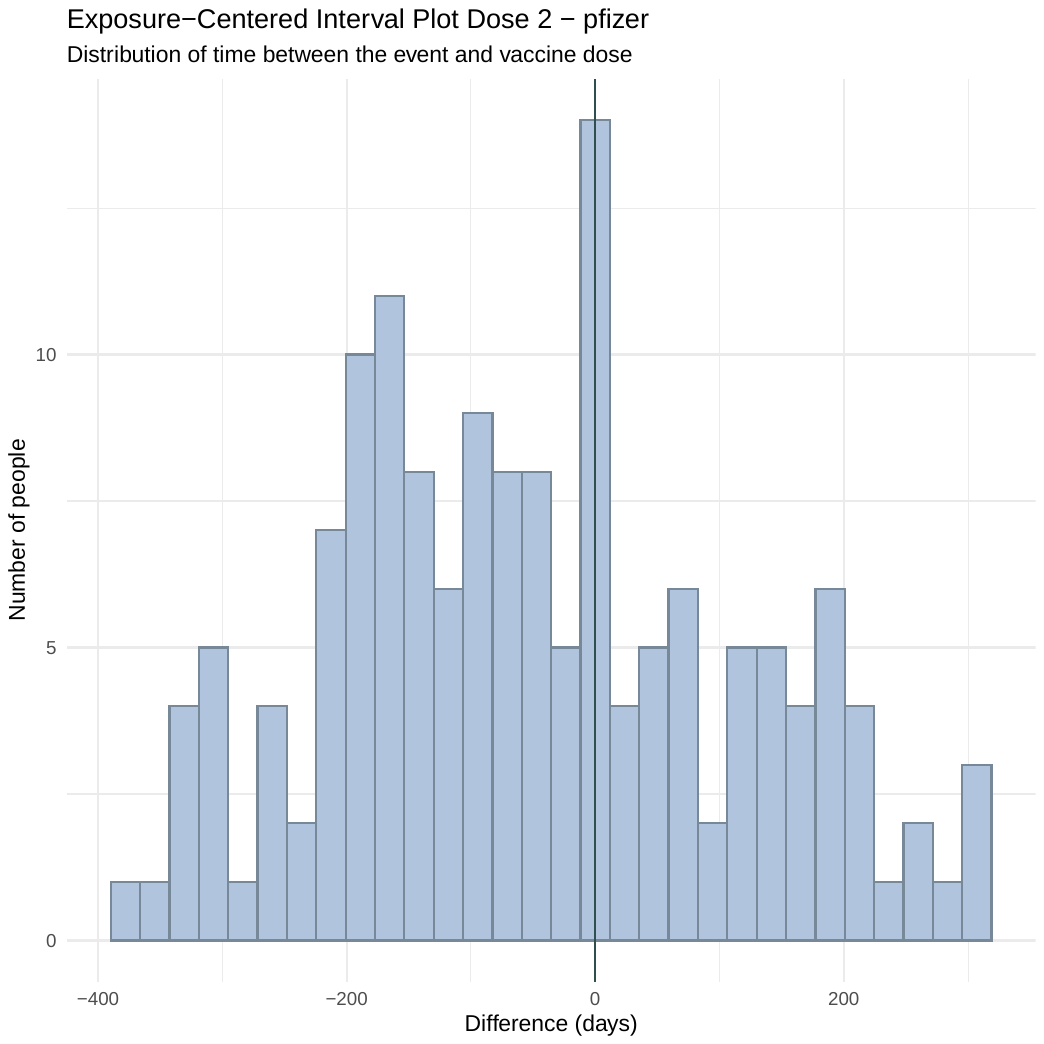

***Figure 3. CPRD Pfizer Dose 1 and 2***

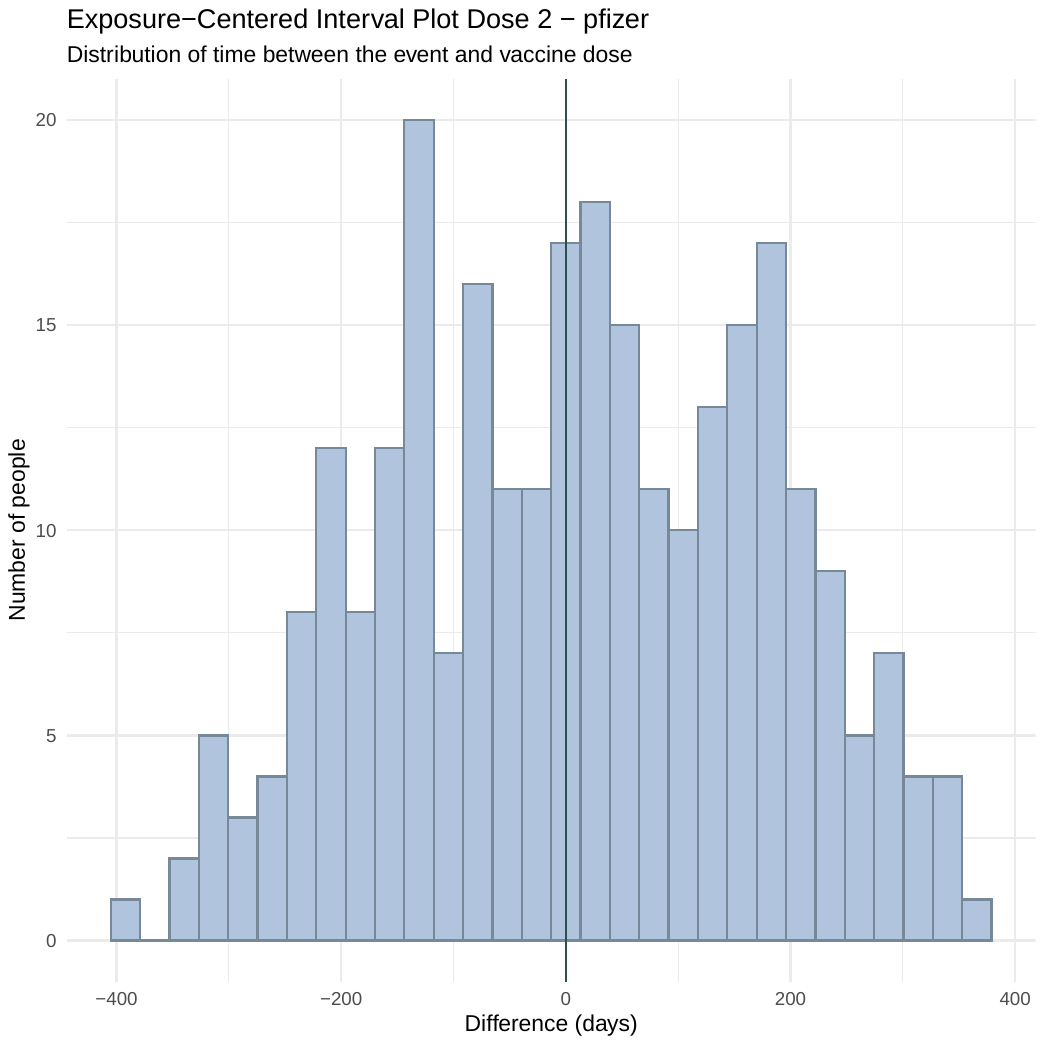

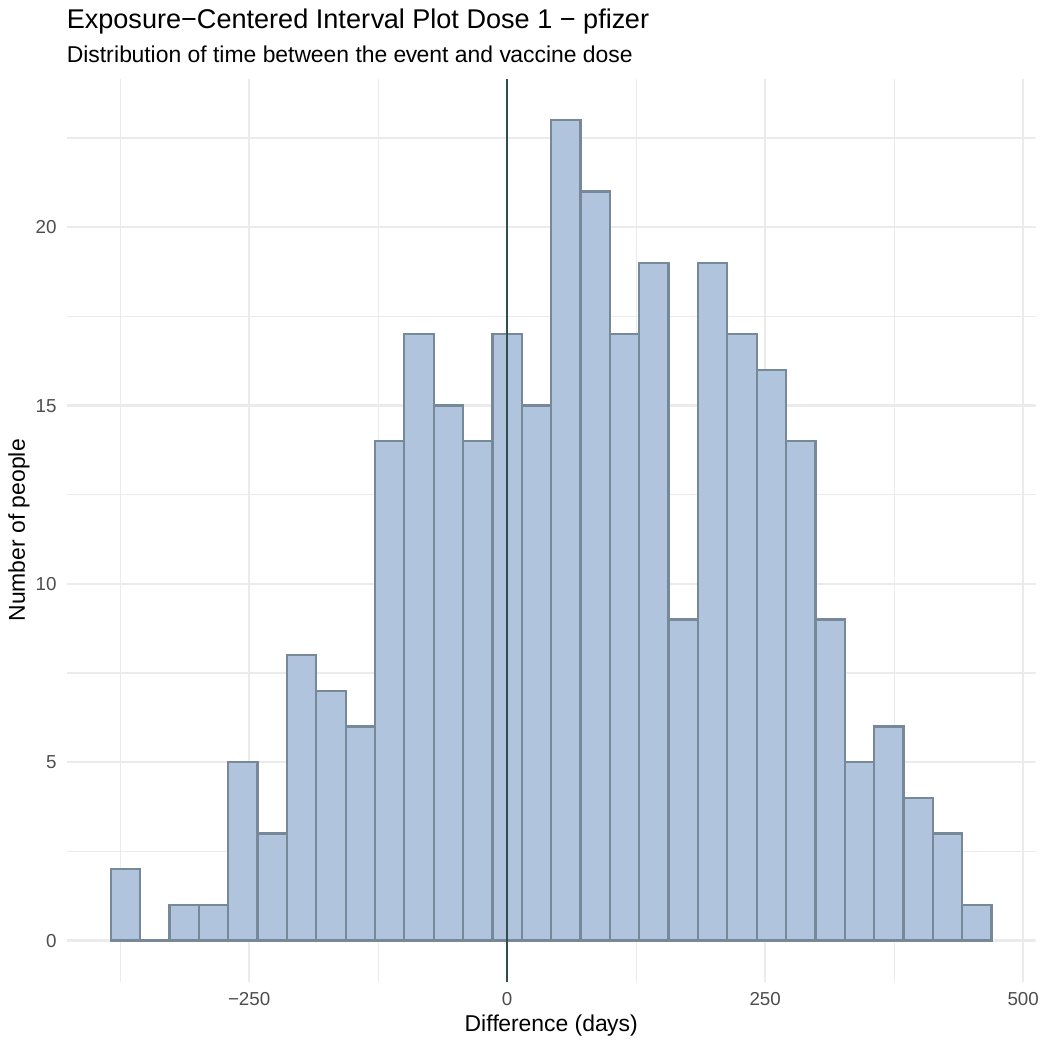

***Figure 4. CPRD AstraZeneca Dose 1 and 2***

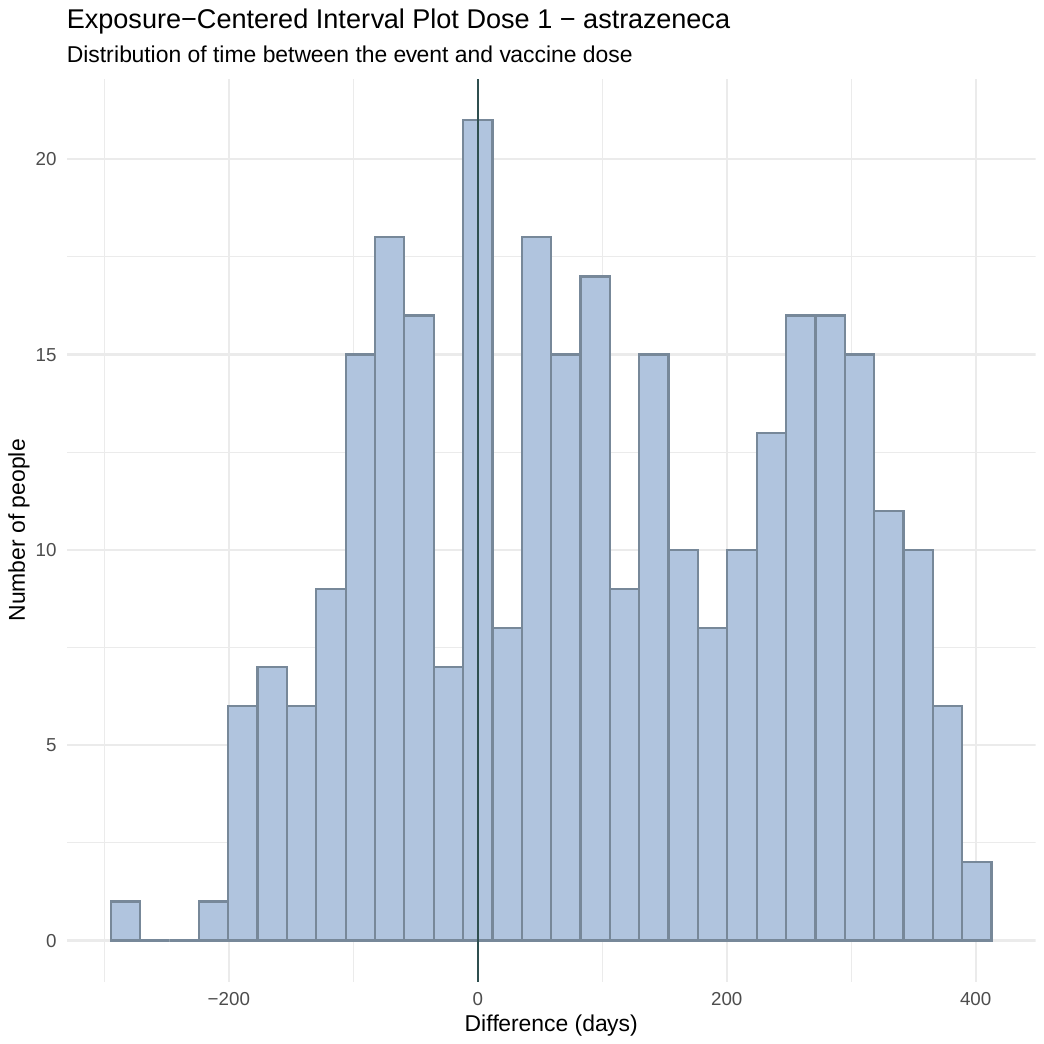

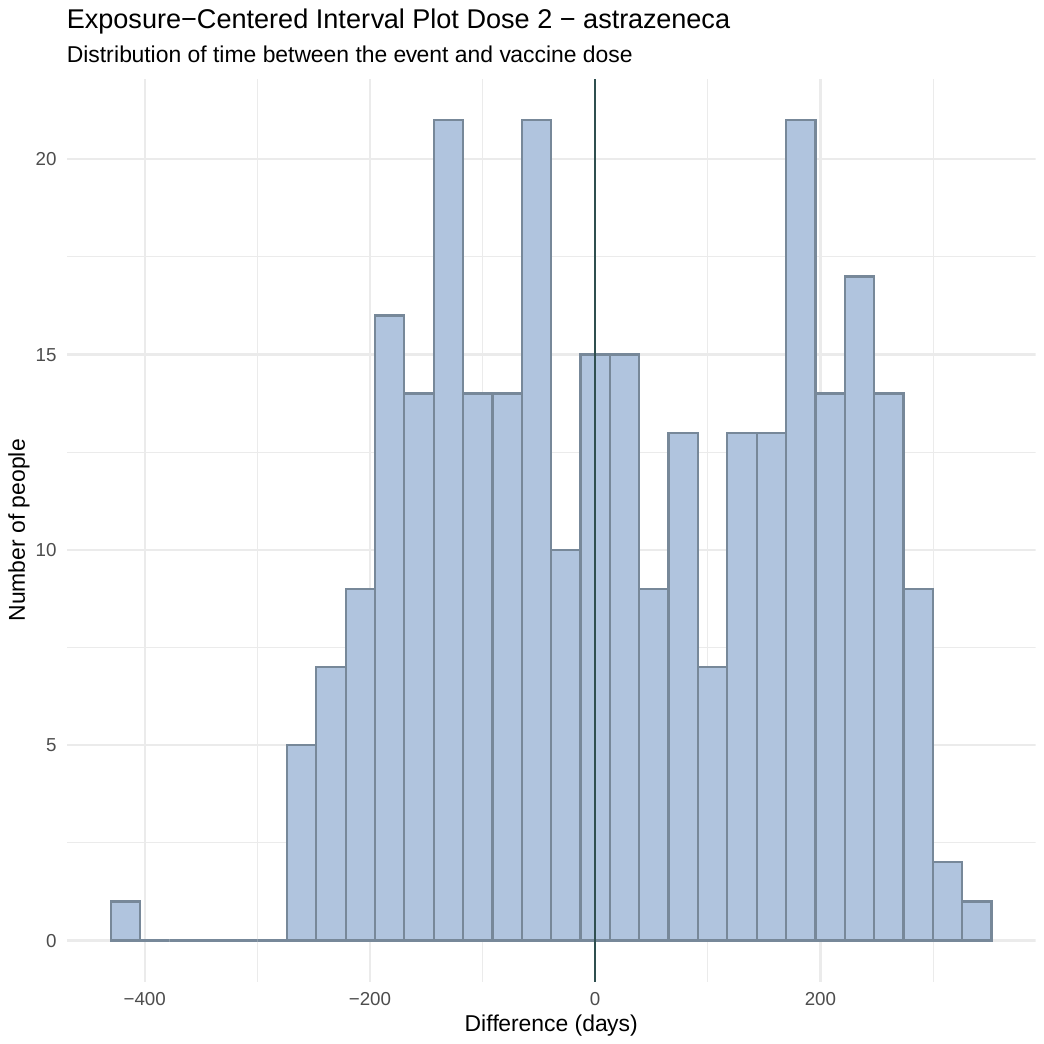
